# Elite neutralizers of human cytomegalovirus are characterized by high magnitude plasma IgG responses against multiple glycoprotein complexes

**DOI:** 10.1101/2022.06.06.22275103

**Authors:** Melissa J. Harnois, Maria Dennis, Dagmar Stöhr, Sarah M. Valencia, Nicole Rodgers, Eleanor C. Semmes, Helen S. Webster, Jennifer A. Jenks, Richard Barfield, Justin Pollara, Cliburn Chan, Christian Sinzger, Sallie R. Permar

## Abstract

**Background:** Human cytomegalovirus (HCMV) is the most common infectious complication of organ transplantation and cause of birth defects worldwide. There are limited therapeutic options and no licensed vaccine to prevent HCMV infection or disease. To inform development of HCMV antibody-based interventions, a previous study identified individuals with potent and broad plasma HCMV-neutralizing activity, termed elite neutralizers (EN), from a cohort of HCMV-seropositive (SP) blood donors. Yet, the specificities and functions of plasma antibodies associated with EN status remained undefined.

**Methods:** We sought to determine the plasma antibody specificities, breadth, and Fc-mediated antibody effector functions associated with the most potent HCMV-neutralizing responses in plasma from EN (n=25) relative to SP (n=19). We measured antibody binding against various HCMV strains and glycoprotein targets, and evaluated Fc-mediated effector functions, antibody dependent cellular cytotoxicity (ADCC) and antibody dependent cellular phagocytosis (ADCP).

**Results:** We demonstrate that elite HCMV neutralizers have elevated IgG binding responses against multiple viral glycoproteins, relative to SP. Our study also revealed potent HCMV-specific ADCC and ADCP activity of EN plasma.

**Conclusions:** We conclude that antibody responses against multiple glycoprotein specificities may be needed to achieve potent plasma neutralization and that potently HCMV elite-neutralizing plasma antibodies can also mediate polyfunctional responses.

## BACKGROUND

Human cytomegalovirus (HCMV) is a ubiquitous beta-herpesvirus that persists as a lifelong infection in human hosts. [1-3] While primary HCMV infection is usually asymptomatic in healthy people, it is the most common viral cause of birth defects worldwide and can cause severe clinical disease and life-threatening complications in immunocompromised individuals. [1, 4-6] Despite ongoing research efforts, no licensed vaccine exists to prevent HCMV infection and current antiviral treatment options remain suboptimal due to long-term toxicities.

Cellular immunity has long been considered the main factor responsible for controlling HCMV infection, but there is evidence that the humoral immune response also serves a critical role in protection. [7-9] Neutralizing antibodies are thought to be especially important for preventing viral entry into host cells as they bind cell-free virus and block interactions between viral glycoproteins and host receptors. [10] More recently, non-neutralizing effector functions, like antibody dependent cellular phagocytosis (ADCP) and antibody dependent cellular cytotoxicity (ADCC), have been highlighted as potentially protective against HCMV infection. [11, 12]

Glycoprotein B (gB) has been a central focus of HCMV vaccine efforts due to its high abundance, immunogenicity, and critical role in facilitating viral entry into host cells. Five distinct genotypes of gB have been described in the literature to date with approximately 95% conserved amino acid identity. [13] The two main sites of sequence variation between gB genotypes are the furin cleavage site and gB antigenic domain 2 (AD2). [14, 15] Within AD2, there are two binding sites; site 1 (AD2S1) is highly conserved and targeted by neutralizing antibodies and site 2 is a variable, non-neutralizing site. [14] While gB is known to induce a strong antibody response, the most potently neutralizing monoclonal antibodies isolated from HCMV-infected individuals target the pentameric complex (PC). [16, 17] The PC is required for viral entry into epithelial, endothelial, and myeloid lineage cells, but not for entry into fibroblasts. [18] A dimer of glycoproteins H and L (gH/gL) exists as part of the PC, but also associates with other glycoproteins, including gO and gB. [19, 20] Antibodies that are specific for the gH/gL and gH/gL/gO complexes have been shown to block HCMV infection in vitro, suggesting that they may also be protective [21]. Yet, it is unknown whether individuals with potent HCMV-neutralizing responses have a dominant neutralizing B cell lineage response against a single glycoprotein complex, like those that develop against neutralizing epitopes on the HIV envelope in HIV-infected individuals with broad neutralizing activity [22], or a combined response against multiple glycoprotein specificities.

Importantly, high titers of HCMV-specific antibodies elicited by natural infection alone are insufficient for preventing re-infection and transmission. HCMV hyperimmune globulin (HIG) is a concentrated antibody product containing pooled plasma from adult seropositive individuals with high-binding HCMV-specific IgG. [16, 23-25] HCMV-HIG has been evaluated for its ability to prevent congenital transmission and infection but has not been established as efficacious in randomized clinical trials. [26-29] Yet, the most efficacious HCMV vaccine to date (gB/MF59) provided approximately 50% protection against HCMV acquisition in adolescent and postpartum women, as well as a reduction in the duration of HCMV viremia and treatment with antiviral therapies in transplant recipients. [6, 30-32] This vaccine elicited high gB-binding IgG titers and non-neutralizing responses, such as virion phagocytosis, but elicited poor neutralization responses and antibodies with limited ability to neutralize heterologous HCMV strains. [11, 33] Moreover, IgG binding to gB expressed on the cell surface was identified as a potential immune correlate of protection associated with the risk of infection across two gB/MF59 vaccine trials, indicating the importance of glycoprotein conformation in effective antibody immunity. [34] There is considerable interest in advancing our understanding of protective anti-HCMV antibody specificities in natural infection to guide vaccine and therapeutic antibody development.

With the goal of improving antibody-based therapeutics, a previous study identified “elite neutralizers” (EN) of HCMV from seropositive blood donors. [35] Plasma from these individuals was selected based on significantly enhanced capacity to neutralize HCMV in a strain independent manner against infection of fibroblasts and endothelial cells, relative to HCMV-HIG and plasma from other HCMV seropositive individuals. [35] The differences in HCMV neutralization capacity of EN versus that of HCMV-seropositive (SP) individuals prompted our investigation into the anti-HCMV antibody glycoprotein binding strength, breadth, specificities, and ability to mediate non-neutralizing effector functions in individuals who develop the most potent neutralizing responses in plasma. The goal of our study was to identify the specificities and characteristics of plasma antibody responses associated with elite neutralizing capacity to guide HCMV vaccine design and the development of more efficacious therapeutic antibody products.

## MATERIALS AND METHODS

See <u>Supplementary Materials and Methods</u> for experimental details.

### Study Population

We obtained 57 plasma samples from previously defined elite HCMV neutralizers (EN; n=25), non-elite neutralizer HCMV seropositive (SP; n=19), and HCMV seronegative (SN; n=13) blood donors. Samples were provided by Christian Sinzger from Ulm University in Germany (Duke University IRB Pro00105640) and were previously obtained from the German Red Cross Blood-Transfusion Service, Baden-Württemberg and Hessen, with informed consent (Ethical Board of Ulm University vote number 53/14). [35]

### Cell culture and virus production

Human monocyte (THP-1) cells were obtained from ATCC and cultured according to ATCC guidelines. HCMV strains TB40/E, AD169r, and Towne were grown on human retinal pigment epithelial (ARPE) cells or human foreskin fibroblast (HFF) cells as previously described [36].

### HCMV Neutralization Assay

The 50% neutralizing titer (NT_50_) was determined for EN and SP against seven strains of HCMV as previously described. [35] HCMV Towne, AD169, TB40/E, VHL/E, Merlin, VR1814, and Toledo strains were used in this neutralization assay. [37]

### Binding Antibody Multiplex Assay (BAMA)

Antibody binding responses against HCMV antigens (gB Domain I, gB Domain II, gB Domain I + II, gB Ectodomain, Pentameric Complex, gH/gL, and gH/gL/gO) were measured by multiplex ELISA as previously described. [34, 36, 38] Antigens were covalently coupled to fluorescent polystyrene beads (Luminex) and incubated with plasma samples. HCMV-specific IgG was detected using phycoerythrin-conjugated goat-anti-human IgG secondary antibody (2 µg/mL, Southern Biotech). Results were acquired on a Bio-Plex 200 system (Bio-Rad) and reported as mean fluorescence intensity.

### IgG Binding and Avidity Enzyme Linked Immunosorbent Assays (ELISA)

Plasma IgG and IgM binding to HCMV whole virions, five gB genotypes (gB1, gB2, gB3, gB4, and gB5), and gB AD2S1 were measured by ELISA as previously described. [36] Briefly, 384-well plates were coated with virus at an optimized concentration of plaque-forming units (PFU) per well (TB40/E, 100 PFU; AD169r, 2700 PFU; Towne, 360 PFU) or with 2 μg/ml of protein per well. Data were collected via SpectroMax and reported as area under the curve (AUC) because full sigmoidal curves were not achieved by all samples. Avidity was reported as Relative Avidity Index (RAI), calculated as the ratio of the OD450 of wells treated for 5 minutes with 7M urea to that of paired 1xPBS-treated wells.

### gB-Transfected Cell Binding Assay (gB-TCB)

Plasma IgG binding to gB expressed on the cell surface was measured as previously described. [34] Human embryonic kidney (HEK)-293T cells were co-transfected with DNA plasmids expressing GFP and gB open reading frame (HCMV Towne) and incubated at 37°C with diluted (1:2500) plasma samples. Cells were stained with Live/Dead Fixable Near-IR Dead Cell Stain (1:1000), followed by PE-conjugated goat-anti-human IgG Fc (1:200), and fixed with 10% formalin prior to acquisition via high throughput sampler (HTS) on the flow cytometer (BD Biosciences). The frequency of PE+ cells was reported for each sample based on the live, singlet, GFP^+^ population.

### Whole HCMV Virion Phagocytosis (ADCP)

AD169r virions (1×10^6^ PFU) were conjugated to AF647 NHS ester prior to incubation with diluted plasma samples (1:100). Virus-antibody immune complexes were then added to THP-1 cells for spinoculation and incubation. Cells were stained with Aqua Live/Dead (1:1000), fixed with 10% formalin, and washed prior to acquisition on the flow cytometer (BD Biosciences) using the HTS. The percentage of AF647^+^ cells was reported for each sample based on the live, singlet population.

### Natural killer (NK) cell CD107a degranulation ADCC assay

Cell-surface expression of CD107a was used as a marker for NK cell degranulation [39, 40] similar to previously described [11, 41]. Live primary human NK cells were added to wells containing AD169 derivative BadrUL131-Y4-GFP-infected MRC-5 cell monolayers. Diluted plasma samples (1:75) were added with Brefeldin A (GolgiPlug, 1 μl/mL, BD), monensin (GolgiStop, 4μl/6mL, BD), and anti-CD107a-FITC (BD, clone H4A3). After a 6-hour incubation, NK cells were washed and stained with a viability dye, anti-CD56-PE/Cy7 (BD, clone NCAM16.2), and anti-CD16-PacBlue (BD, clone 3G8). [11] The frequencies of CD107a+ live NK cells were determined by flow cytometry. Final data represent specific activity, determined by subtraction of non-specific activity observed in assays performed with mock-infected cells.

### Statistical Analysis

The statistical analysis plan was created prior to analysis. Raw data were internally reviewed prior to analysis, based on established quality control criteria for each assay.

Differences in the antibody binding responses between groups were analyzed via Fisher Exact Test or Wilcoxon Rank Sum Test. For analyses where a cutoff was established, data below cutoff (defined as mean of the seronegatives plus 3 standard deviations) was set to cutoff prior to analysis. For the gB binding breadth analysis, established cutoffs were used to determine binding for each sample to each genotype. Differences in the number of gB genotypes bound per sample between EN and SP groups were determined by Fisher Exact Test. In the analysis of HCMV neutralization, we first log transformed the data and fit a Tobit regression model [42, 43] (a censored linear regression model allowing for data at lower limit of detection) while adjusting for HCMV-specific IgG to assess differences between EN and SP. For ADCP, we logit transformed the data prior to analysis. Binding responses were converted to antibody concentration (µg/ml) in GraphPad Prism. Plasma IgG binding responses were then normalized to HCMV-specific IgG concentration and Wilcoxon Rank Sum Tests were performed to assess differences between EN and SP. Multiple testing correction was performed per set of analysis using Benjamini-Hochberg false discovery rate (FDR) adjustment. [44] Statistical significance was pre-defined as p<0.05, with a FDR-adjusted p-value of less than 0.2, reflecting the hypothesis-generating nature of this study. [34, 45] All statistical analyses were performed within R.

## RESULTS

### EN and SP plasma IgG and IgM binding responses to HCMV

To determine differences between EN and SP antibody responses against HCMV, plasma IgG binding to TB40/E, AD169r, and Towne whole virus was measured by ELISA. These viruses were selected based on glycoprotein B genotype and differences in PC expression (listed in Table 1). EN plasma exhibited significantly higher IgG binding to all tested strains, relative to SP controls (all p<0.0001) (Fig 1A). Next, we explored whether EN and SP individuals differed in terms of time since primary infection or re-infection by measuring HCMV-specific plasma IgG avidity and IgM binding responses. EN exhibited a significant increase in IgG avidity for AD169r (p=0.004), but no statistically significant differences in avidity for TB40/E (p=0.09) or Towne (p=0.07), relative to SP controls (Fig 1B). Additionally, while the proportion of EN with detectable plasma HCMV TB40E-specific IgM was higher than that of SP (78.6.0% vs 21.4%, respectively), this difference was not statistically significant as determined by Fisher Exact Test (FDR p-value=0.07) (Fig 1C).

**Table 1.** HCMV neutralization potency after adjustment for HCMV-specific IgG concentration. Results from the Tobit regression are shown in log scale as non-elite seropositive (SP) controls compared to elite neutralizers (EN). Statistical significance was defined as p<0.05 and a multiple testing adjusted p-value less than 0.2. ^a^ Estimates of the log difference between groups adjusting for IgG concentration if the outcome were uncensored.

| HCMV Strain | gB genotype<br>[13, 50] | PC | Log Difference <sup>a</sup> | p-value | FDR p-value |
| --- | --- | --- | --- | --- | --- |
| Toledo | gB3 | - | -3.64 | <0.0001 | <0.0001 |
| TB40/E | gB1 | Intact | -3.47 | <0.0001 | <0.0001 |
| VR1814 | gB3 | Intact | -3.66 | <0.0001 | <0.0001 |
| VHL-E | gB1 | Intact | -1.87 | <0.0001 | <0.0001 |
| Towne | gB1 | - | -1.25 | <0.0001 | <0.0001 |
| AD169 | gB2 | - | -2.29 | 0.0002 | 0.0002 |
| Merlin | gB1 | Intact | -0.75 | 0.42 | 0.42 |

**Figure 1.**
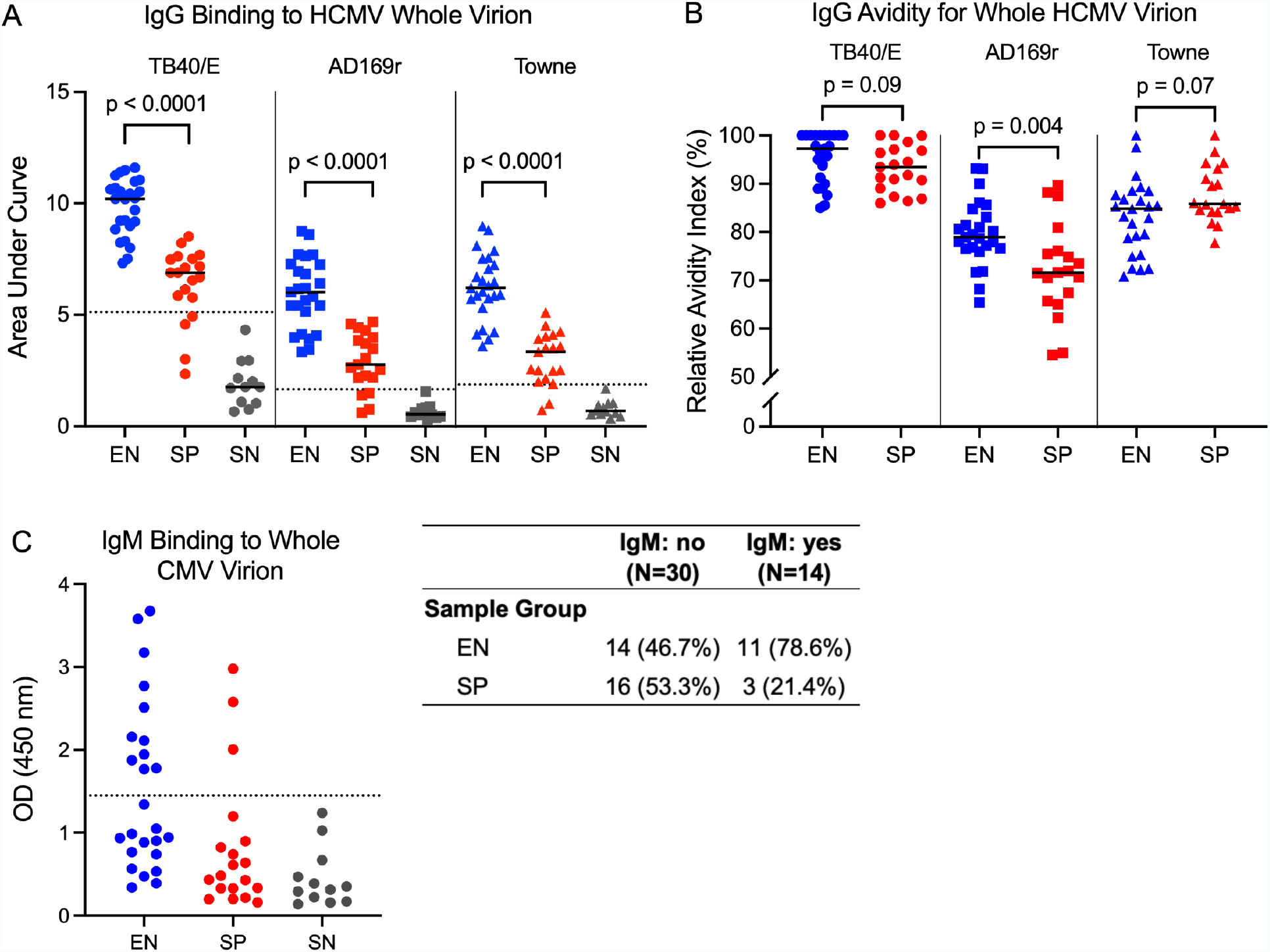
Plasma IgG and IgM binding responses to whole HCMV virions in elite neutralizers compared to seropositive and seronegative individuals. **(A)** Area under the curve (AUC) for IgG binding to TB40/E (median AUC: EN=10.21, SP=6.89), AD169r (median AUC: EN=6.02, SP=2.77), and Towne (median AUC: EN=6.22, SP=3.35) whole virus by enzyme-linked immunosorbent assay (ELISA). AUC median is indicated. **(B)** Relative IgG binding avidity index (RAI) to TB40/E (median RAI: EN=97.3, SP=93.5; p=0.0938), AD169r (median RAI: EN=79.0, SP=71.6; p=0.00392), and Towne (median RAI: EN=84.9, SP=85.8; p=0.0695) whole virus by ELISA. RAI median is indicated. **(C)** Optical density (OD) at 450nm showing EN and SP plasma with TB40/E-specific IgM binding. All samples were run in duplicate. n = 25 EN, n = 19 SP, and n=13 SN; blue symbols, elite neutralizers (EN); red symbols, seropositive controls (SP); grey symbols, seronegative controls (SN). Symbols: circle = TB40/E, square = AD169r, triangle = Towne. Wilcoxon Rank Sum Test (A,B) and Fisher Exact Test (C) were used to calculate statistical differences between groups; statistical significance was defined as p<0.05 and a multiple testing adjusted p-value less than 0.2. All reported p-values are FDR adjusted. Dotted lines indicate the negative cutoff, determined by the average SN binding response plus 3x the standard deviation.

### EN and SP plasma IgG binding to HCMV surface glycoproteins and peptides

To investigate plasma IgG binding responses to known HCMV glycoprotein targets of anti-HCMV antibodies, we employed a binding antibody multiplex assay (BAMA). EN plasma IgG bound with statistically significantly higher magnitudes to all glycoprotein complexes tested (all p<0.0001), including gH/gL dimer, gH/gL/gO trimeric complex, PC, and gB ectodomain, relative to SP plasma (Fig. 2A and 2B). A similar trend was observed for plasma IgG binding to gB Domains among EN, including gB Domain I, Domain II, Domain I + II, cell-associated gB, and gB AD2S1, relative to SP (all p<0.0001) (Fig. 2B-D).

**Figure 2.**
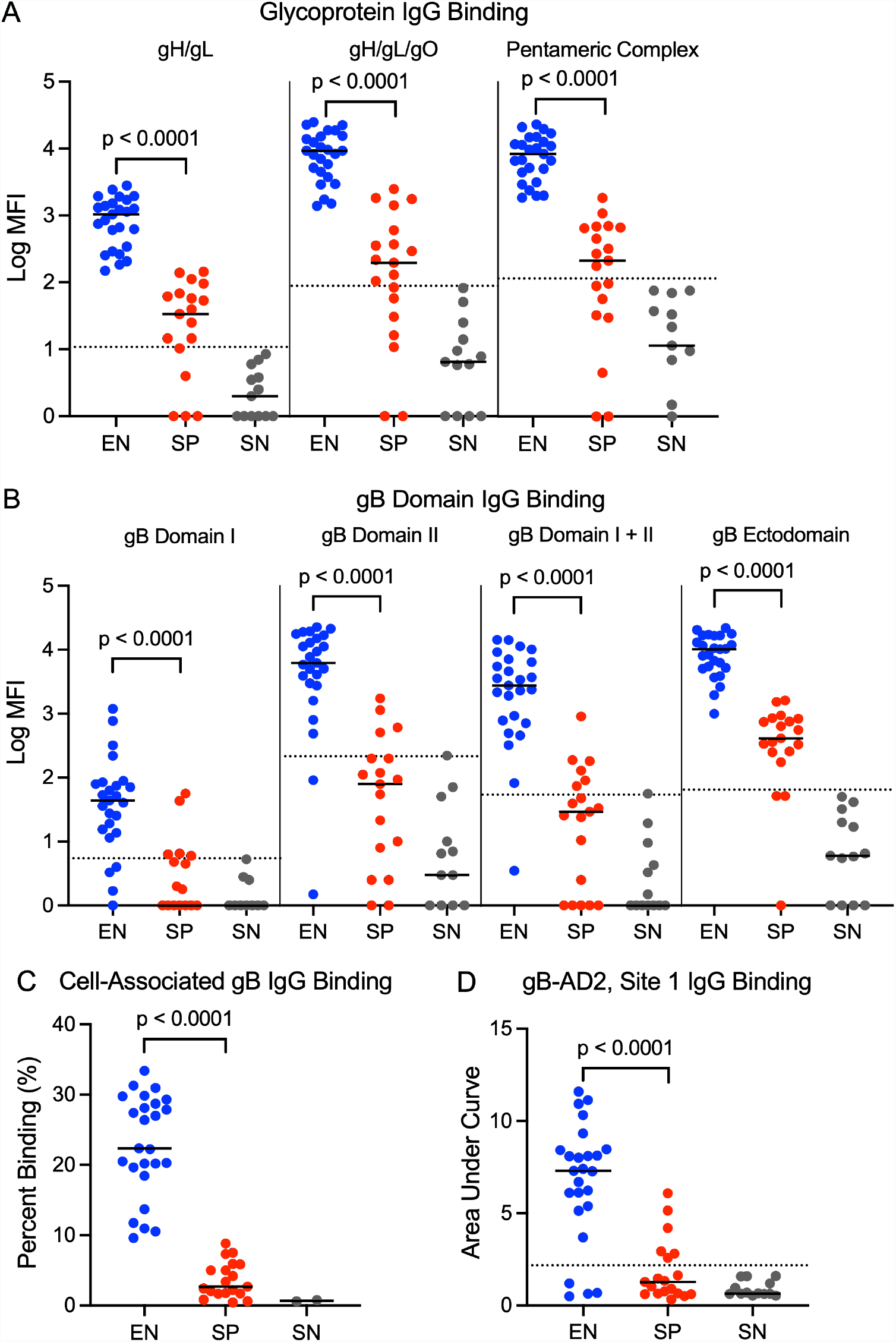
HCMV glycoprotein conformational and linear epitope IgG binding in HCMV elite neutralizers compared to seropositive and seronegative individuals. Mean fluorescence intensity (MFI) of plasma IgG binding to **(A)** HCMV surface glycoproteins: gH/gL (median log_10_MFI: EN=3.02, SP=1.53), gH/gL/gO (median log_10_MFI: EN=3.97, SP=2.29), Pentameric Complex (PC) (median log_10_MFI: EN=3.92, SP=2.33); **(B)** HCMV gB Domains: gB Domain I (median log_10_ MFI: EN=1.64, SP=0.763), gB Domain II (median log_10_ MFI: EN=3.79, SP=2.33), gB Domains I + II (median log_10_ MFI: EN=3.44, SP=1.73), and gB ectodomain (median log_10_MFI: EN=4.01, SP= 2.61). **(C)** Frequency of plasma IgG binding to cell-associated gB (median percent binding: EN=26.4, SP=3.38). **(D)** Area under curve for plasma IgG binding to known neutralizing epitope, gB AD2S1 (median AUC: EN=7.30, SP=2.18). Samples were run in duplicate. n = 25 EN, n = 19 SP, and n=13 SN; blue symbols, elite neutralizers (EN); red symbols, seropositive controls (SP); grey symbols, seronegative controls (SN). Wilcoxon Rank Sum Test was used to calculate statistical differences between groups; statistical significance was defined as p<0.05 and a multiple testing adjusted p-value less than 0.2. All reported p-values are FDR adjusted. Dotted lines indicate the negative cutoff, determined by the average SN binding response plus 3x the standard deviation.

### EN and SP plasma IgG binding magnitude and breadth across HCMV gB genotypes

HCMV gB variants are classified into five common gB genotypes (gB1 through gB5). Previous studies have reported elevated risk of HCMV disease among individuals with multi-gB genotype HCMV infections, demonstrating potential implications for future vaccine candidates that incorporate multiple gB genotypes, especially given the moderate efficacy of a single valent gB protein antigen vaccine. [6, 13, 31, 32, 46] Thus, we wanted to determine if there were detectable differences in the breadth of gB-specific IgG binding responses between EN and SP plasma. We define binding breadth as the ability of antibodies from a single donor to bind multiple genotypes of the same glycoprotein antigen; in this case we evaluated plasma IgG binding across 5 discrete gB genotypes by ELISA. EN exhibited a significantly higher magnitude of binding to all 5 gB genotypes (all p<0.0001) relative to SP controls (Fig. 3 A and B). We then assessed plasma IgG binding breadth by calculating the number of gB genotypes bound by each plasma sample using established cutoffs. There was no significant difference in IgG binding breadth across gB genotypes between EN and SP groups (p = 0.07).

**Figure 3.**
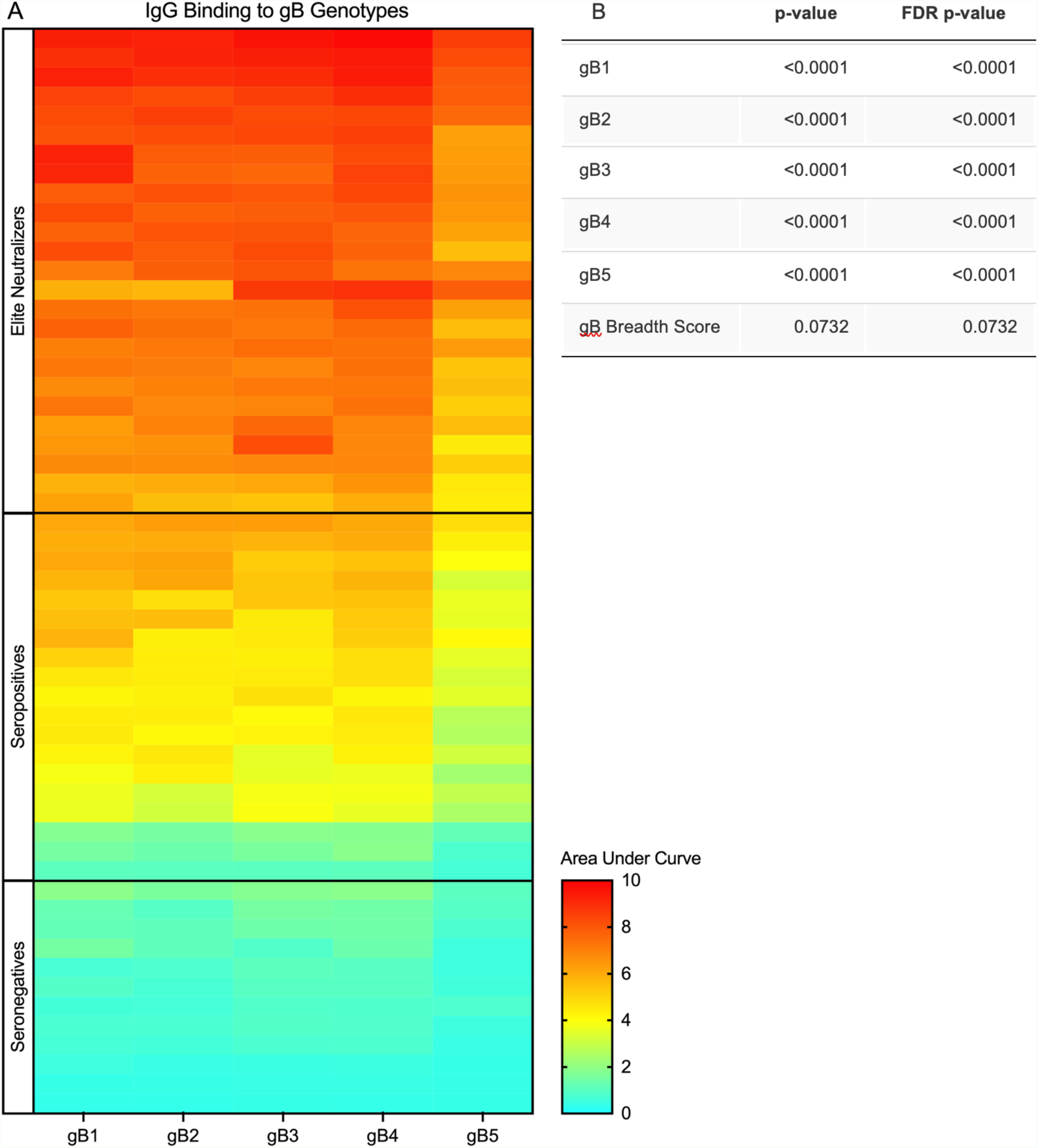
Plasma IgG binding to multiple gB genotypes in HCMV elite neutralizers, seropositive, and seronegative individuals. **(A)** Area under the curve (AUC) for IgG binding to multiple glycoprotein B (gB) genotypes: gB1, gB2, gB3, gB4, gB5 as measured by ELISA. n = 25 EN, n = 19 SP, n=13 SN. **(B)** Wilcoxon Rank Sum Test was used to test for statistical differences between the magnitude of IgG binding to each gB genotype across groups; Fisher Exact Test was used to calculate statistical differences in gB binding breadth (number of gB genotypes with value above the seronegative cutoff) between groups. Significance was defined as p<0.05 and a multiple testing adjusted p-value less than 0.2.

### Relationship between total HCMV-specific IgG levels, neutralization responses, and glycoprotein-specific IgG binding

Previous work by Falk et. al demonstrated that plasma anti-HCMV IgG titer does not determine HCMV neutralizing status. [37] To follow up on this finding in the present cohort, we first interpolated the HCMV-specific plasma IgG concentration for each sample based on the HCMV-HIG standard binding curve against TB40/E. A Tobit regression model was applied to the neutralization data from this cohort to adjust for the impact of HCMV-specific IgG concentration on EN and SP plasma neutralization capacity. The model confirmed that plasma neutralization response magnitude against most tested HCMV strains was still associated with EN and SP status (Table 1) after adjusting for total HCMV-specific plasma IgG concentration. This indicates that the quality of the antibody response, and not just the magnitude of the total HCMV-specific IgG response, is important in determining plasma neutralization potency. The exception to this observation was the Merlin strain of HCMV, for which the Tobit regression revealed no distinction between EN and SP after adjustment for HCMV-specific IgG concentration. Next, we normalized the glycoprotein-specific plasma IgG binding responses to the total HCMV-specific IgG concentration and performed a Wilcoxon Rank Sum Test on the normalized outputs. After normalization, the magnitude of plasma HCMV glycoprotein-specific IgG binding remained significantly higher among EN relative to that of SP to all measured targets except for gB genotype 2 (p=0.36) (Table 2).

**Table 2.**
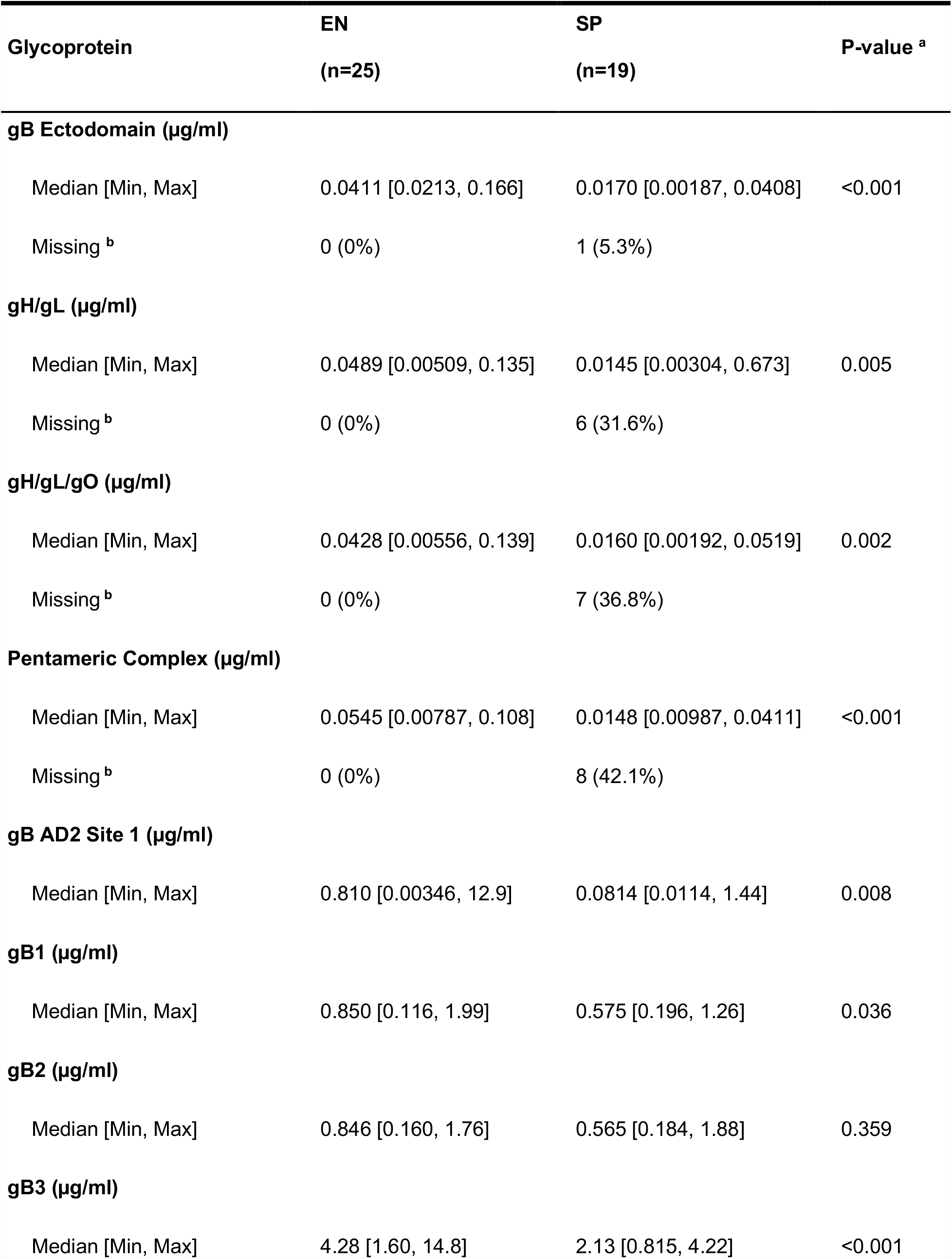
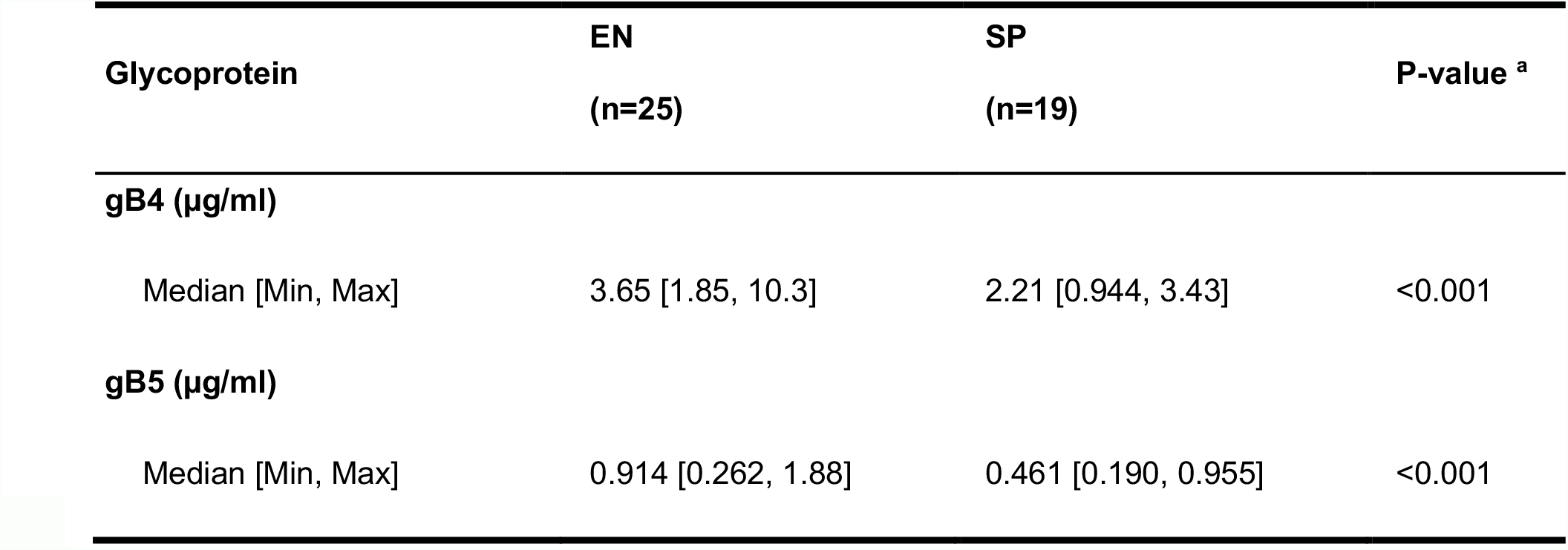
HCMV glycoprotein-specific IgG concentration normalized to total HCMV-specific IgG concentration. ^**a**^ P-value from Wilcoxon-Rank Sum test. ^**b**^ unable to interpolate concentration.

| **Glycoprotein** | **EN**<br>**(n=25)** | **SP**<br>**(n=19)** | **P-value <sup>a</sup>** |
| --- | --- | --- | --- |
| **gB Ectodomain (µg/ml)** |  |  |  |
| Median [Min, Max] | 0.0411 [0.0213, 0.166] | 0.0170 [0.00187, 0.0408] | <0.001 |
| Missing <sup>b</sup> | 0 (0%) | 1 (5.3%) |  |
| **gH/gL (µg/ml)** |  |  |  |
| Median [Min, Max] | 0.0489 [0.00509, 0.135] | 0.0145 [0.00304, 0.673] | 0.005 |
| Missing <sup>b</sup> | 0 (0%) | 6 (31.6%) |  |
| **gH/gL/gO (µg/ml)** |  |  |  |
| Median [Min, Max] | 0.0428 [0.00556, 0.139] | 0.0160 [0.00192, 0.0519] | 0.002 |
| Missing <sup>b</sup> | 0 (0%) | 7 (36.8%) |  |
| **Pentameric Complex (µg/ml)** |  |  |  |
| Median [Min, Max] | 0.0545 [0.00787, 0.108] | 0.0148 [0.00987, 0.0411] | <0.001 |
| Missing <sup>b</sup> | 0 (0%) | 8 (42.1%) |  |
| **gB AD2 Site 1 (µg/ml)** |  |  |  |
| Median [Min, Max] | 0.810 [0.00346, 12.9] | 0.0814 [0.0114, 1.44] | 0.008 |
| **gB1 (µg/ml)** |  |  |  |
| Median [Min, Max] | 0.850 [0.116, 1.99] | 0.575 [0.196, 1.26] | 0.036 |
| **gB2 (µg/ml)** |  |  |  |
| Median [Min, Max] | 0.846 [0.160, 1.76] | 0.565 [0.184, 1.88] | 0.359 |
| **gB3 (µg/ml)** |  |  |  |
| Median [Min, Max] | 4.28 [1.60, 14.8] | 2.13 [0.815, 4.22] | <0.001 |

| Glycoprotein | EN<br>(n=25) | SP<br>(n=19) | P-value <sup>a</sup> |
| --- | --- | --- | --- |
| **gB4 (µg/ml)** |  |  |  |
| Median [Min, Max] | 3.65 [1.85, 10.3] | 2.21 [0.944, 3.43] | <0.001 |
| **gB5 (µg/ml)** |  |  |  |
| Median [Min, Max] | 0.914 [0.262, 1.88] | 0.461 [0.190, 0.955] | <0.001 |

### Investigation of non-neutralizing antibody effector functions in EN and SP plasma

Aside from neutralizing responses, Fc-mediated antibody effector functions also potentially contribute to protection from HCMV infection [41]. Thus, we compared the ability of EN and SP plasma to promote HCMV-specific ADCP and ADCC in flow-based assays using whole HCMV virions and HCMV-infected fibroblasts, respectively. Using a linear regression model, we determined that EN plasma exhibited significantly enhanced ability to promote ADCP of the whole HCMV virion and ADCC responses against infected cells, relative to SP (p<0.0001 and p=0.01, respectively) (Fig. 4A and 4B). However, after adjusting for total HCMV-specific IgG concentration in the linear regression model, the ADCC and ADCP responses did not differ between EN and SP groups (Table 3).

**Table 3.** Non-neutralizing HCMV antibody responses after adjustment for HCMV-specific IgG concentration. Results from linear regression after adjustment for total HCMV specific IgG concentration are shown in log scale as non-elite seropositive (SP) controls compared to elite neutralizers (EN). Statistical significance was defined as p<0.05 and a multiple testing adjusted p-value of less than 0.2. ^a^ Estimates of the log difference between groups adjusting for IgG concentration.

| HCMV Strain | gB genotype [13] | PC | Log Difference <sup>a</sup> | p-value |
| --- | --- | --- | --- | --- |
| TB40/E | gB1 | Intact | -0.306 | 0.232 |
| AD169r | gB2 | Repaired | -0.158 | 0.509 |
| Towne | gB1 | - | 0.959 | 0.742 |

**Figure 4.**
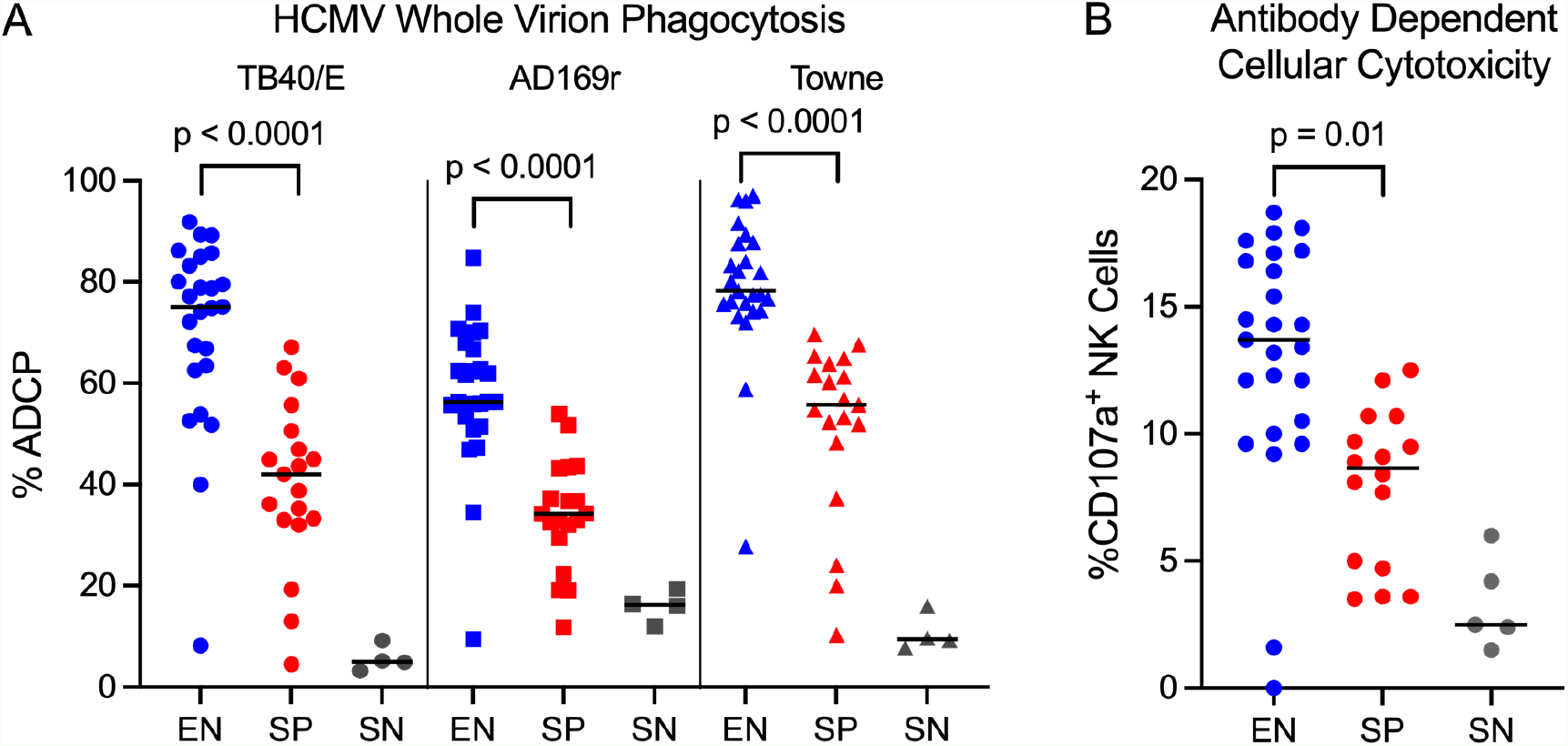
Plasma IgG non-neutralizing effector function activity in HCMV elite neutralizers, seropositive, and seronegative individuals. **(A)** Percentage of AF647+ THP-1 cells were measured to quantify antibody-mediated phagocytosis of TB40/E (median %ADCP: EN=75.1, SP=42.1), AD169r (median %ADCP: EN=56.3, SP=34.2), and Towne (median %ADCP: EN=78.3, SP=55.8) whole HCMV virions. **(B)** Percentage of CD107a+ NK cells were measured to quantify plasma antibody-mediated cellular cytotoxicity against HCMV-infected cells (median %CD107a+ NK cells: EN=13.7, SP=8.65). n = 25 EN, n = 19 SP, n=5 SN.

## DISCUSSION

In this study, we found that EN plasma can mediate potent HCMV-specific polyfunctional responses, including neutralizing and non-neutralizing effector functions. We observed significantly elevated HCMV-specific IgG binding responses to three strains of HCMV and higher avidity for AD169r among EN plasma, relative to that of SP. Moreover, we found that EN plasma exhibited significantly higher magnitudes of IgG binding to all tested HCMV glycoproteins known to be targets of neutralizing antibodies, namely gB, PC, gH/gL, and gH/gL/gO. These findings suggest that robust IgG responses to multiple glycoproteins may be critical to achieving potently neutralizing antibody responses against HCMV, as opposed to single-specificity IgG potency driving EN status. This is notably distinct from what is reported in studies of potent HIV-specific neutralizing responses in HIV-infected individuals, wherein certain plasma IgG epitope specificities, such as the HIV envelope CD4 binding site, are dominant in elite neutralizers. [22, 47] Thus, a live-attenuated viral vaccine formulation with an intact PC and/or a vaccine with multiple HCMV glycoprotein immunogens, as is being pursued currently in a phase 3 trial by Moderna (NCT05085366), may be needed to elicit the most potent plasma neutralization across cell types and provide robust protection against HCMV infection.

We observed that EN plasma IgG bound with significantly higher magnitude to gB Domain II and AD2S1, known neutralizing epitopes, and to gB Domain I, which is targeted by neutralizing and non-neutralizing antibodies. [33, 48] Relative to SP, EN plasma also exhibited significantly increased IgG binding magnitudes across the five gB genotypes. However, there were no statistically significant differences in IgG binding breadth across gB genotypes between groups. After normalizing HCMV glycoprotein-specific IgG concentrations by total HCMV-specific IgG concentration, we observed that EN plasma maintains significantly elevated binding responses against all targets, except for gB genotype 2, relative to SP. Interestingly, our correlative analysis of normalized responses revealed a negative correlation between plasma IgG against neutralizing epitope gB AD2S1 and gB genotype 2, but only among EN (Supplementary Figure 2). One potential explanation is that although AD2S1 is known to be highly conserved across genotypes, sequence variations in nearby residues may conformationally shield AD2S1 in gB genotype 2 and disrupt antibody binding to this site and that, relative to SP, a larger proportion of gB-specific antibodies in EN plasma is directed towards this neutralizing epitope. Moreover, the differences observed between EN and SP binding to cell-associated gB could indicate that EN plasma contains antibodies specific for unique epitopes of gB that are exposed when gB is associated with the cell surface, which was previously identified as a correlate of protection in the gB/MF59 vaccine trials [34]. Overall, our findings indicate that incorporating multiple glycoprotein B genotypes into future vaccine formulations may not be necessary to confer broad and potent neutralizing responses to HCMV, whereas including multiple glycoprotein antigens with appropriate conformations to allow antibody access to neutralizing epitopes could be more impactful on elicitation of broad and potent neutralizing responses.

Finally, we observed that EN plasma exhibits significantly elevated neutralization against HCMV relative to SP, even after adjusting for total HCMV-specific IgG concentration. Thus, neutralizing status does not appear to be determined solely by the magnitude of the HCMV-specific antibody titer, but rather by the quality and specificities of HCMV-specific antibodies generated in response to infection. Interestingly, this was not observed for the HCMV Merlin (gB1) strain, which has been characterized as potentially more representative of clinical isolates than some lab-adapted strains [49]. Yet, while EN plasma was also capable of mediating significantly higher magnitudes of ADCP and ADCC relative to SP, these responses did not distinguish between groups after adjusting for HCMV-specific IgG in a linear regression model. This suggests that unlike the distinct quality of HCMV-neutralizing responses between EN and SP individuals, differences in ADCP and ADCC response magnitudes between groups are due to the higher HCMV-specific IgG concentration in EN plasma, not a distinction in antibody response quality.

There are several limitations to this study. First, the sample size is relatively low, and all plasma samples were selected from blood donors at a single timepoint. This study was designed as a cross-sectional study, so our results cannot definitively confirm acute versus chronic infection within the cohort. Interestingly, while the proportion of HCMV-specific IgM responses was higher in EN, the difference was not statistically significant between groups. We cannot rule out that IgM responses are contributing to EN status, but our IgM binding and IgG avidity data suggest that time since primary infection or re-infection is not distinct between EN and SP. Therefore, the higher neutralization capacity observed among EN plasma is likely not solely attributable to a recent primary HCMV infection. Additionally, due to limited sample volume, we were unable to assess the relative contributions of domain-specific IgG responses towards the potent and broad neutralizing capacity of EN plasma through glycoprotein-specific IgG depletion studies. Further investigation is required to understand these aspects of HCMV elite neutralizer plasma IgG responses.

This study represents the first characterization of neutralizing antibody responses to multiple HCMV surface glycoproteins, including multiple gB glycoproteins and conformations, elicited by natural infection that contribute to elite HCMV neutralization. Overall, our data reveal that plasma or monoclonal antibodies from EN individuals can mediate strong polyfunctional antibody responses against HCMV, suggesting that the potency and efficacy of currently available HCMV-antibody therapeutics, such as HCMV-HIG, may be improved by selecting for elite HCMV neutralizing plasma donors. HCMV is capable of infecting various cell types via distinct entry mechanisms and uses multiple glycoprotein complexes to do so, making it highly unlikely that targeting a single epitope or even a single glycoprotein will be sufficient to elicit a protective response across multiple strains of HCMV. Thus, our results further support the development of vaccines that incorporate multiple glycoprotein complexes needed for infection of various cell types to elicit broad and potent neutralizing antibodies and polyfunctional antibody responses against HCMV.

## Data Availability

All data produced in the present study are available upon reasonable request to the authors.

## Acknowledgements

We thank Jessica Falk who initiated this collaboration together with SRP at the IHW 2018 in Vancouver. We wish to acknowledge support from the Biostatistics, Epidemiology and Research Design (BERD) Methods Core at Duke University, funded through Grant Award Number UL1TR002553 from the National Center for Advancing Translational Sciences (NCATS), a component of the NIH. This work was also supported by the Translating Duke Health Initiative (awarded to CC, RB), the Duke Center for Human Systems Immunology, the Else Kröner-Fresenius-Foundation (awarded to CS, 2016-A126), T32 Training Grant (awarded to MJH, 2T32AI052077-16A1), Medearis CMV Scholar Award (awarded to MJH), and NIH National Institute of Allergy and Infectious Disease P01 (awarded to SRP, 3P01AI129859).

## Meetings

This work was previously presented in part at the 2021 International Herpesvirus Workshop, Virtual Conference (poster).

