## Supplementary Figures and Materials for "Elite neutralizers of human cytomegalovirus are characterized by high magnitude plasma IgG responses against multiple glycoprotein complexes"

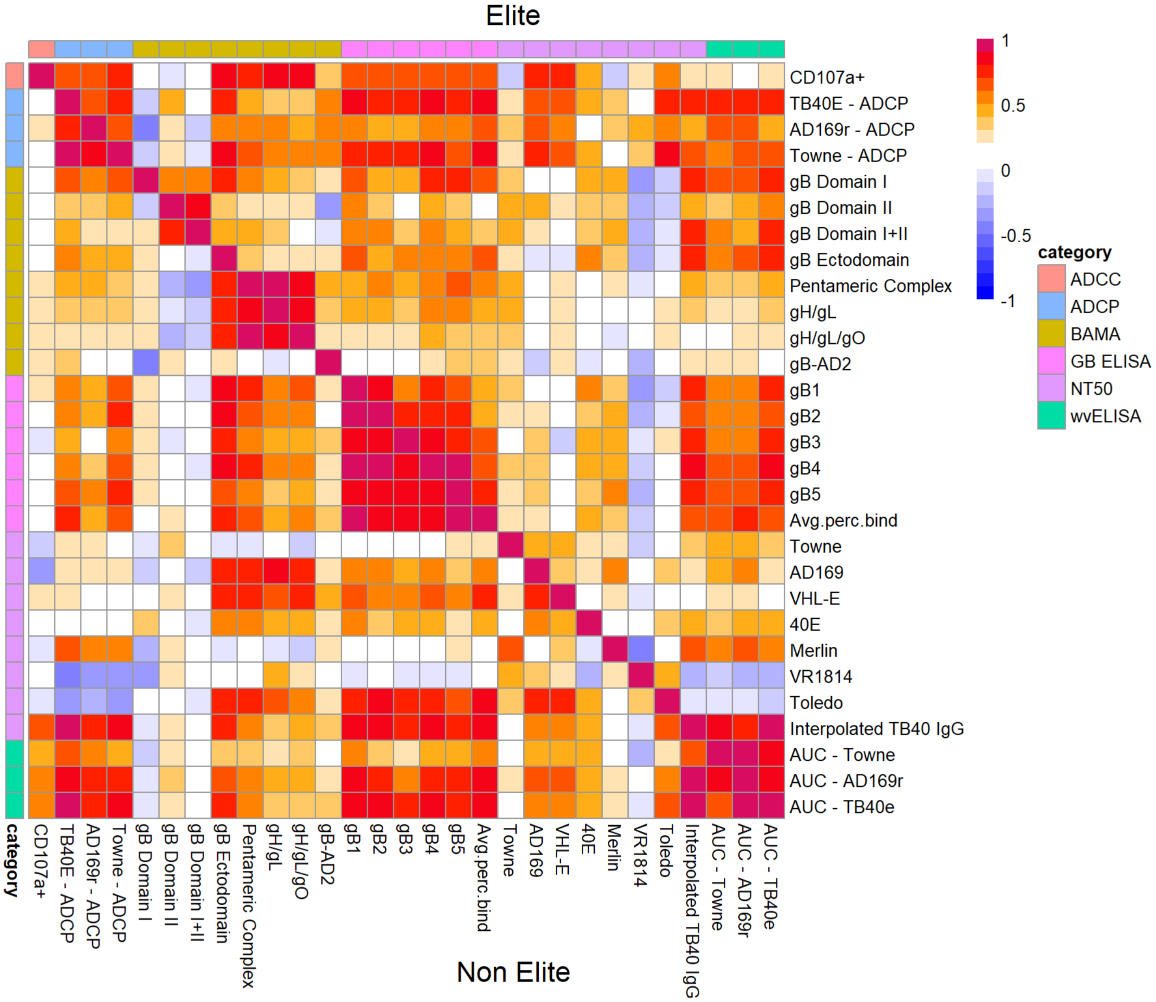


**Supplementary Figure 1.** Correlative analysis between all measured binding and functional responses separated by neutralizing status.

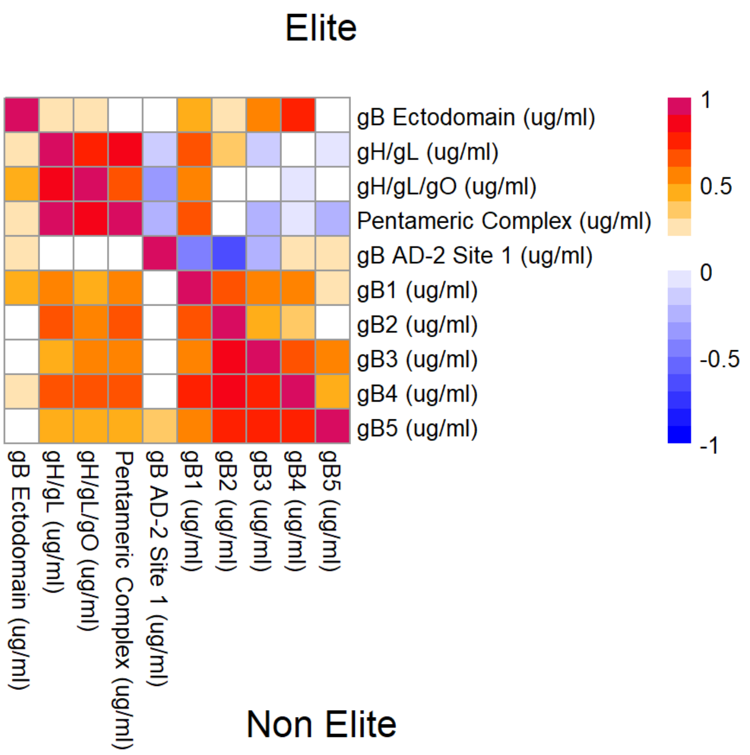


**Supplementary Figure 2.** Correlative analysis of binding responses after adjustment for total HCMV-specific IgG concentration, separated by neutralizing status.

### SUPPLEMENTARY MATERIALS AND METHODS

*Study Population.* We obtained 57 plasma samples from previously defined elite HCMV neutralizers (EN; n=25), non-elite neutralizer HCMV seropositive (SP; n=19), and HCMV seronegative (SN; n=13) blood donors. Samples were provided by Christian Sinzger from Ulm University in Germany (Duke University IRB Pro000105640) and were previously obtained from the German Red Cross Blood-Transfusion Service, Baden-Württemberg and Hessen, with informed consent (Ethical Board of Ulm University vote number 53/14). [35] Using a two-step screening approach, plasma samples were previously identified and categorized based on their ability to broadly and potently neutralize HCMV in vitro. [37] More specifically, elite neutralizing plasma was defined as the top 2.5% of neutralizers against TB40/E infection in both fibroblasts and endothelial cells and had the ability to neutralize seven different strains of HCMV. [35] Low neutralizers were defined as plasma samples that fell below 97.5$^{th}$ percentile of neutralizing ability relative to reference plasma. [35]

*HCMV Neutralization Assay.* The 50% neutralizing titer ($NT_{50}$) was determined for EN and SP against seven strains of HCMV as previously described. [35] Briefly, human foreskin fibroblast (HFF) cells were infected with one of seven HCMV strains pre-incubated with serially diluted plasma. Cells were stained with an HCMV immediate early antigen-specific monoclonal antibody (Clone E13, Argene Biosoft) and a horseradish peroxidase (HRP)-conjugated IgG secondary antibody (Dako). Readout was measured as the optical density at 492 nm ($OD_{492}$). HCMV strains that were used in this neutralization assay included Towne, AD169, TB40/E, VHL/E, Merlin, VR1814, and Toledo.

*Binding Antibody Multiplex Assay (BAMA).* Antibody responses against purified HCMV antigens were investigated by a multiplex ELISA. Antigens of interest included gB Domain I, gB Domain II, gB Domain I + II, gB Ectodomain, Pentameric Complex, gH/gL, and gH/gL/gO. [34, 36, 38] These antigens were covalently coupled to fluorescent polystyrene beads (Luminex). HCMV-seropositive plasma samples were diluted in assay diluent (phosphate-buffered saline, 5% normal goat serum, 0.05% Tween 20, and 1% Blotto milk) and run in duplicate at a 1:500 dilution. Diluted plasma was incubated for 30 minutes with the antigen-coupled beads. Samples were then washed with wash buffer (0.1% BSA, 0.02% Tween in 1X phosphate buffer saline) and incubated with phycoerythrin-conjugated goat-anti human IgG for 30 minutes (2 μg/mL, Southern Biotech). Beads were washed a third time with wash buffer and mean fluorescence intensity (MFI) was acquired on a Bio-Plex 200 Plate Reader (Bio-Rad). Cytogam (CSL Behring) was used as a positive control to confirm consistent assay performance across runs and HCMV-seronegative plasma samples were included as negative controls. Blank beads were used as a negative control to account for nonspecific binding and blank wells were used to assess assay background. Preset assay criteria required a coefficient of variation less than or equal to 20% and greater than or equal to 100 beads counted per sample. Negative cutoffs were established for each antigen based on the average MFI of seronegative samples plus 3x the standard deviation, while MFI > 23,000 was considered 'maximum' response.

*Whole Virus Enzyme Linked Immunosorbent Assay (WV ELISA).* Plasma IgG binding to and avidity for HCMV Towne, AD169r, and TB40/E strains were measured by ELISA. First, 384-well plates were coated with each virus at optimized PFU (TB40/E, 100 PFU/well; AD169r, 2700 PFU/well; Towne, 360 PFU/well). Viruses were diluted in 0.1 M sodium bicarbonate buffer, pH 9.6. Coated plates were incubated at 4ºC overnight. Plates were washed one time with wash buffer (95% DI water, 4% 25X PBS, 1% Tween-20). Plates were then blocked with blocking buffer (4% whey, 15% goat serum, 0.5% tween 20, 80.5% 1X PBS) and incubated at either 4ºC overnight or for a minimum of 2 hours at RT. During the incubation, samples and standard controls were diluted 1:30 in blocking buffer and then serially diluted 1:3 in a 96-well plate. After blocking, plates were washed, and diluted samples and controls were added to the plate in duplicate. Plates were incubated again for 2 hours at RT. After the incubation, plates were washed two times and 7M urea was added to half of the wells, according to the plate map. Plates were incubated for 5 minutes at RT. Secondary antibody (Goat Anti-Human IgG-HRP, Jackson ImmunoResearch) was prepared at a 1:5000 dilution in blocking buffer. After washing two times, secondary antibody was added to each well. Plates were sealed and incubated for 1 hour at RT. Plates were then washed four times and room temperature SureBlue Reserve TMB Substrate (VWR) was added to each well. Plates were incubated for 10 minutes while shielded from light. After the incubation, TMB Stop Solution (VWR) was added to each well. Plates were immediately read at 450 nm via SpectroMax using the SoftMax software interface. Results are reported as area under the curve (AUC) because full sigmoidal curves were not achieved by all samples. Cytogam (CSL Behring) was used as a positive control to ensure consistent performance across runs and HCMV-seronegative plasma samples were included as negative controls. Negative cutoffs were established for each virus based on the average AUC of HCMV seronegative samples plus 3x the standard deviation.

*IgM WV ELISA.* Plasma IgM binding to TB40/E was measured by ELISA. First, the virus was used at an optimal concentration (100 PFU/well) to coat 384-well plates. Virus was diluted with 0.1 M sodium bicarbonate buffer, pH 9.6. Coated plates were incubated at 4ºC overnight. Plates were washed one time with wash buffer (95% DI water, 4% 25X PBS, 1% Tween-20), blocked with blocking buffer (4% whey, 15% goat serum, 0.5% tween 20, 80.5% 1X PBS) and incubated at either 4ºC overnight or for a minimum of 2 hours at RT. During the incubation, samples and standard controls were diluted 1:20 in blocking buffer as a single point dilution. Previously characterized IgM-positive human plasma samples were used as positive controls and HCMV-seronegative samples were included as negative controls. After blocking, plates were washed and diluted samples and controls were added to the plate in duplicate. Plates were incubated again for 2 hours at RT. Secondary antibody (Goat Anti-Human IgM-HRP, Jackson

ImmunoResearch) was prepared at a 1:5000 dilution in blocking buffer. Plates were washed two times and diluted secondary antibody was added to each well. Plates were sealed and incubated for 1 hour at RT. Plates were then washed four times and room temperature SureBlue Reserve TMB Substrate (VWR) was added to each well. Plates were incubated for 10 minutes while shielded from light. After the incubation, TMB Stop Solution (VWR) was added to each well. Plates were immediately read at 450 nm via SpectroMax using the SoftMax software interface. Negative cutoffs were established based on the average OD450 of HCMV seronegative samples plus 3x the standard deviation.

*gB-Transfected Cell Binding Assay (gB-TCB).* Human embryonic kidney (HEK)-293T cells were grown to approximately 50% confluency in a T75 flask. Cells were co-transfected using the Effectene Transfection Reagent Kit (Qiagen) with DNA plasmids expressing green fluorescent protein (GFP) and plasmids expressing full length gB open reading frame from the Towne strain (Sino Biological). Transfected cells were incubated for 48 hours at 37ºC, 5% $CO_2$ and the presence of successfully transfected cells was confirmed visually by GFP expression under a fluorescence microscope. Cells were then dissociated from the flask by incubation with TrypLE (Gibco) and a full media exchange was performed into HEK293T growth media (DMEM, 10% FBS, 2.5% HEPES, 1% Pen/Strep). The live cell count was used to measure 200,000 cells/well into a 96-well U-bottom plate (Corning). Plates were centrifuged for 5 minutes at 500 x g and cells were resuspended in Human TruStain Fc Block (BioLegend) diluted 1:1000 in wash buffer (PBS + 0.1% FBS). After a 5-minute incubation at RT, plasma samples were diluted 1:2500 in HEK-293T growth media. HCMV immunoglobulin standard (Cytogam, CSL Behring) was included as a positive control. Blank wells and HCMV-seronegative plasma samples were included as negative controls. Plates were centrifuged for 5 minutes at 500 x g and cells were resuspended in diluted plasma. Plates were incubated for 2 hours at 37ºC, 5% $CO_2$. Cells were then washed and stained with Live/Dead Fixable Near-IR Dead Cell Stain (Invitrogen) at a 1:1000 dilution for

20 minutes at RT. Cells were washed and stained for 25 minutes at 4ºC with PE-conjugated goat anti-human IgG Fc (Southern Biotech) diluted at 1:200. Cells were washed and fixed with 10% Formalin for 10 minutes at RT. After one final wash, cells were resuspended in wash buffer prior to acquisition on the flow cytometer (BD Biosciences). The percentage of PE+ cells was reported for each sample based on the live, singlet, GFP+ population.

*gB binding breadth ELISA*. Plasma IgG binding to five gB genotypes (gB1, gB2, gB3, gB4, and gB5) as well as to glycoprotein B Antigenic Domain 2, Site 1 (AD2S1) of HCMV were measured by ELISA. First, 384-well plates were coated with 2 µg/ml of protein diluted in 0.1 M sodium bicarbonate buffer, pH 9.6. Coated plates were incubated at 4ºC overnight. Plates were then washed one time with wash buffer (95% DI water, 4% 25X PBS, 1% Tween-20), blocked with blocking buffer (4% whey, 15% goat serum, 0.5% tween 20, 80.5% 1X PBS), and incubated at 4ºC overnight. For gB genotype ELISAs, samples and standard controls were diluted 1:100 in blocking buffer and serially diluted 1:3 in a 96-well plate. For AD2S1 ELISA, samples and standard controls were diluted 1:30 in blocking buffer and serially diluted 1:4 in a 96-well plate. Cytogam was used as a positive control (CSL Behring) and HCMV-seronegative samples were used as negative controls. After the blocking step, plates were washed, and diluted samples and controls were added to the 384-well plate in duplicate. Plates were incubated for 2 hours at RT. Secondary antibody (Goat Anti-Human IgG-HRP, Jackson ImmunoResearch) was prepared at a 1:5000 dilution in blocking buffer. After the incubation, plates were washed two times and secondary antibody was added to each well. Plates were sealed and incubated for 1 hour at RT. Plates were then washed four times and room temperature SureBlue Reserve TMB Substrate (VWR) was added to each well. Plates were incubated for 6 minutes while shielded from light. After the incubation, TMB Stop Solution (VWR) was added to each well. Plates were immediately read at 450 nm on the SpectroMax using the SoftMax software interface. Results are reported as area under the curve (AUC) because full sigmoidal curves were not achieved by all samples. Negative cutoffs were established for each virus based on the average area under the curve (AUC) of seronegative samples plus 3x the standard deviation.

*Whole HCMV Virion Phagocytosis (ADCP).* AD169r virions (1x$10^6$ PFU) were conjugated to AF647 NHS ester (Invitrogen) and diluted 1:10 in wash buffer (1x PBS + 0.1% FBS). Patient plasma samples were diluted 1:100 in wash buffer. Cytogam was used as a positive plate control, while known seronegatives, a non-specific mAb, and AD169r-AF647-only wells (no antibody) were included as negative controls for the assay. Diluted sera and controls (10 ul) were transferred to round bottom 96-well plates in duplicate. Next, 10 ul of conjugated virus (AD169r-AF647) was added to each well. Plates were covered and incubated for 2 hours at 37ºC. During this incubation, THP-1 cells were harvested, counted, and resuspended in fresh growth media (RPMI + 10%FBS) at a concentration of 250,000 cells/ml. After the sera and virus incubation, 50,000 THP-1 cells were added to each well of the round bottom 96-well plates containing sera and virus. Plates were centrifuged at 1200xg at 4ºC for 1 hour and then incubated at 37ºC for 1 hour. Plates were then spun at 1200xg for 5 minutes. Media was removed and 100 ul of Aqua live/dead stain was added at a 1:1000 dilution for 15 minutes. Cells were spun again and washed 1x with wash buffer. Cells were fixed with 100 ul 10% formalin for 15 minutes and finally resuspended in 100 ul PBS. Cells were acquired on the BD Fortessa using the high throughput sampler (HTS) input.

*Natural killer (NK) cell CD107a degranulation ADCC assay***.** Cell-surface expression of CD107a was used as a marker for NK cell degranulation [39, 40] similar to previously described [11, 41]. Briefly, target cells (MRC-5 fibroblasts) were plated at 3.5x$10^4$ cells/well in a 96-well plate and infected with the AD169 derivative BadrUL131-Y4-GFP at an MOI of 1.0 for 48 hours. Effector cells were primary human NK cells isolated by negative selection with magnetic beads (Human NK cell isolation kit, Miltenyi Biotech) from peripheral blood mononuclear cells (PBMC) of a healthy donor after overnight rest in cell culture media supplemented with 10 ng/mL IL-15. Live NK cells ($3.5x10^4$) were added to each well containing HCMV-infected MRC-5 cell monolayers, and plasma samples were added at a final dilution of 1:75. Brefeldin A (GolgiPlug, 1 µl/mL, BD Biosciences), monensin (GolgiStop, 4µl/6mL, BD Biosciences), and anti-CD107a-FITC (BD Biosciences, clone H4A3) were added to each well and the plates were incubated for 6 hours at 37ºC in a humidified 5% CO2 incubator. NK cells were recovered and stained with Live/Dead Aqua Dead Cell Stain (Molecular Probes, ThermoFisher) followed by anti-CD56-PE/Cy7 (BD Biosciences, clone NCAM16.2), and anti-CD16-PacBlue (BD Biosciences, clone 3G8). Data was acquired using an LSR Fortessa cytometer (BD Biosciences),and analyzed with FlowJo 10 software (BD Biosciences). Data is reported as the frequency of CD107a positive (+) live NK cells (singlets, lymphocytes, aqua blue-, CD56+ and/or CD16+, CD107a+). All final data represent specific activity, determine by subtraction of non-specific activity observed in assays performed with mock-infected cells.

*Statistical Analysis.* The statistical analysis plan was created prior to data analysis. Raw data were internally reviewed prior to analysis, based on established quality control criteria for each assay. Differences in the antibody binding responses between groups were analyzed via Fisher Exact Test or Wilcoxon Rank Sum Test. For analyses where a cutoff was established, data below cutoff (defined as mean of the seronegatives plus 3 standard deviations) was set to cutoff prior to analysis. For the gB binding breadth analysis, established cutoffs were used to determine binding for each sample to each genotype. Differences in the number of gB genotypes bound per sample between EN and SP groups were determined by Fisher Exact Test. In the analysis of HCMV neutralization, we first log transformed the data and fit a Tobit regression model [42, 43] (a censored linear regression model allowing for data at lower limit of detection) while adjusting for HCMV-specific IgG to assess differences between EN and SP. For ADCP, we logit transformed the data prior to analysis. Binding responses were converted to antibody concentration (µg/ml) in

GraphPad Prism. Plasma IgG binding responses were then normalized to HCMV-specific IgG concentration and Wilcoxon Rank Sum Tests were performed to assess differences between EN and SP. Multiple testing correction was performed per set of analysis using Benjamini-Hochberg false discovery rate (FDR) adjustment. [44] Statistical significance was pre-defined as $p<0.05$, with a FDR-adjusted p-value of less than 0.2, reflecting the hypothesis-generating nature of this study. [34, 45] All statistical analyses were performed within R.
